# Morphological Landscape Mapping Decodes Pathological Heterogeneity and Proteomic Programs in HCC

**DOI:** 10.64898/2025.11.27.25341163

**Authors:** Xiaodong Wang, Yifei Cheng, Xulang Peng, Jinda Li, Jing Han, Zhicheng Yang, Guang-Yu Ding, Haichao Zhao, Peixuan Qu, Qingyun Sun, Zichen Chang, Tonghui Zou, Biyu Wang, Tinggui Huang, Zhuosong Cheng, Siqi Guo, Sainan Ma, Jingwen Wang, Chuang Liu, Xiaolei Zhang, Shu Zhang, Long Tian, Henry Han, Lirong Ai, Liming Wang, Xiao-Ying Wang, Jian Zhou, Jia Fan, Qiang Gao, Hu Zhou, Xiyang Liu, Jie-Yi Shi

## Abstract

**Background & Aims:** Intratumour heterogeneity (ITH) drives the clinical trajectory of HCC, yet routine pathology relies on global classifications that often mask local architectural diversity. We developed an unsupervised artificial intelligence framework to deconvolute HCC histology into a comprehensive lexicon of morphotypes, link them to *in situ* proteomic programs, and derive spatial biomarkers for precision prognostication.

**Approach & Results:** We developed Morphological Landscape Mapping (MLM), a deep-learning framework integrating multi-scale clustering with optimal-transport similarity. Applied to 1,448 whole-slide images across four independent HCC cohorts, MLM distilled the tumour landscape into 16 reproducible morphological phenotypes associated with distinct prognostic outcomes. Deep *in situ* proteomics (>8,000 unique proteins) linked MLM-derived morphotypes to specific molecular programs. Notably, an aggressive loss-of-adhesion morphotype exhibited hypoxia signalling activation and focal-adhesion downregulation, consistent with its fragmented architecture, whereas favourable trabecular morphotypes retained xenobiotic metabolic machinery. Building on these phenotypes, we developed the Morphotypic Spatial Entropy Index (MSEI) to quantify architectural complexity within the tumour ecosystem. In multivariable Cox models, MSEI remained an independent predictor of survival in two large HCC cohorts (adjusted HR 2.57, 95% CI: 1.66-3.98; and adjusted HR 2.21, 95% CI 1.28-3.81) after accounting for tumor stage and vascular invasion, and it further stratified risk within early-stage (BCLC 0-A) disease.

**Conclusions:** MLM transforms routine H&E slides into quantitative, molecularly characterized maps of HCC heterogeneity. By coupling morphology with underlying biology and spatial organization, this framework provides a scalable, cost-effective foundation for morphology-guided precision oncology.

## Introduction

Cancer evolution drives intratumour heterogeneity (ITH) at both molecular and morphological levels, fostering therapeutic resistance and unpredictable clinical trajectories^1–5^. Hepatocellular carcinoma (HCC), one of the most heterogeneous malignancies^6^, epitomizes these challenges, with persistently high recurrence rates despite therapeutic advances^7,8^. While early research primarily utilized bulk multi-omics^3,4^, recent efforts have shifted toward spatial omics technologies^9,10^. However, clinical translation of these approaches remains limited by high costs, technical complexities, and poor scalability, which prevent their adoption as routine diagnostic tools^11^.

Fundamentally, molecular heterogeneity manifests as morphological diversity, which is directly observable in routine hematoxylin-and-eosin (H&E)-stained slides—the universal cornerstone of cancer diagnostics^12–15^. However, the full potential of morphological assessment remains untapped due to structural limitations in current classification systems. Standard diagnostics, governed by the World Health Organization (WHO) classification, typically enforce a strict hierarchy where a specific histological subtype is assigned only when a distinct pattern constitutes >50% of the tumour area (or even >80% for subtypes like Clear Cell HCC)^16^. This reductionist approach inherently masks the compositional heterogeneity of HCC, obscuring sub-dominant yet structurally distinct components that may drive aggressive behaviour. This limitation is most acute in the “not otherwise specified” (NOS) category—a heterogeneous “wastebasket” that encompasses over 65% of all HCC cases^17^. We posit that the clinical ambiguity of NOS reflects unresolved heterogeneity, where distinct phenotypic mixtures are concealed by a broad diagnostic label. Consequently, there is a pressing need for computational frameworks capable of deconvoluting this ambiguity into a precise, reproducible spectrum of morphological subtypes.

Artificial intelligence (AI), particularly pathology foundation models, offers a transformative opportunity to decode these complex patterns quantitatively^18–23^. Yet, two critical barriers persist. First, most supervised strategies rely on labour-intensive annotations^24–26^ or predefined labels, limiting unbiased discovery of novel disease-related features^27–29^. In contrast, current unsupervised approaches often struggle to identify robust, spatially consistent phenotypes that are reproducible across diverse patient populations^30,31^. Second, there remains a critical disconnect between visual morphological phenotypes and their underlying molecular drivers. This “black box” nature hampers mechanistic understanding of why specific morphological patterns serve as prognostic biomarkers.

To address these challenges, we developed Morphological Landscape Mapping (MLM), an unsupervised framework applied to 1,448 HCC cases across four independent cohorts (Fig. 1). MLM resolved the visual complexity of HCC into a reproducible lexicon of 16 distinct morphotypes, effectively decoding the latent substructure of NOS tumours. By combining phenotype-guided laser-capture microdissection (LCM) with deep *in situ* proteomics, we bridged morphology and molecular function, revealing the molecular programs that underlie key prognostic morphotypes—from preserved metabolic machinery to hypoxia-associated loss of focal-adhesion components. Moving beyond phenotypic composition to spatial organization, we introduced the Morphotypic Spatial Entropy Index (MSEI) to quantify the architectural disorder of the tumour ecosystem. This novel biomarker quantifies the architectural disorder of the tumour ecosystem, enabling precise risk stratification within established clinical stages. By unlocking the prognostic and biological information encoded in routine pathology slides, MLM provides a scalable, morphology-guided foundation for precision oncology.

**Figure 1.**
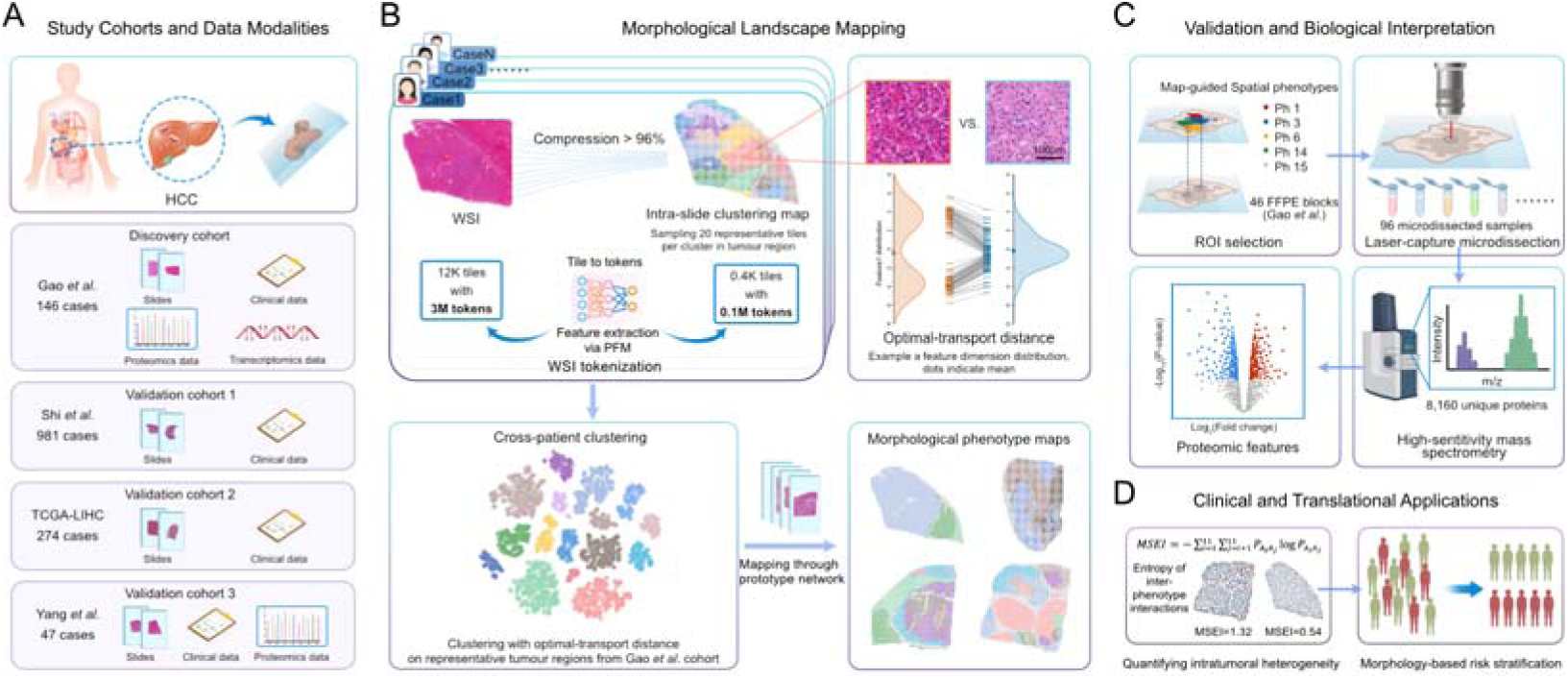
Discovery, validation, and clinical application of morphological phenotypes in hepatocellular carcinoma (HCC). (A) Study cohorts and data modalities. The study leverages four independent HCC cohorts to enable a multi-stage discovery and validation workflow. The discovery cohort (Gao *et al*., n = 146 cases) contains whole-slide images (WSIs), transcriptomic, proteomic, and clinical data. The validation cohorts include Shi *et al*. (n = 981 cases) and TCGA-LIHC (n = 274 cases) which provide WSI and clinical data for independent prognostic verification, and Yang *et al*. cohort (n = 47 cases) which provides bulk proteomic data for molecular validation. (B) A pathology foundation model (PFM) encodes each gigapixel WSI into tens of thousands of tile-level feature tokens. Guided sampling reduces the tokens by >96 % while preserving intratumoural diversity, achieved by intra-slide clustering followed by the selection of 20 representative tiles per subregion. Inter-patient clustering with the Leiden algorithm and an optimal-transport (OT) distance metric then defines cross-patient morphological phenotypes. An OT-Prototype network is trained to recognise these phenotypes and project spatial phenotype maps onto every WSI across cohorts. (C) Biological validation via deep proteomics. Phenotype maps guide laser-capture microdissection (LCM) of specific morphological regions of interest (ROIs) from formalin-fixed, paraffin-embedded (FFPE) tissues. Untargeted mass spectrometry analysis of these samples identified 8,160 unique proteins, confirming that visual phenotypes correspond to distinct molecular signatures. (D) Clinical and translational application. Intratumour heterogeneity (ITH) is quantified using the Morphotypic Spatial Entropy Index (MSEI), a metric derived from the entropy of spatial interaction networks between morphological phenotypes. By capturing the architectural complexity of the tumour ecosystem, MSEI enables precise, morphology-based patient risk stratification.

## Patients and Methods

### Study Design and Patient Cohorts

Our study utilized four independent, retrospective cohorts, totalling 1,448 HCC patients, to systematically discover and validate morpho-molecular signatures (Fig. 1A). The core discovery of morphological phenotypes was performed using a multi-omics cohort (Gao *et al*.^32^, n=146) from the CPTAC project, which provided paired whole-slide images (WSIs) and deep proteomic data. Following discovery, the prognostic value of these phenotypes was assessed in the large-scale Shi *et al*. cohort (n=981) ^29^, and independently validated in the TCGA-LIHC cohort (n=274). To further corroborate the link between morphology and protein expression, a fourth cohort with bulk proteomics data (Yang *et al*.^33^, n=47) was used for external validation. The WSIs for the Shi *et al*. and TCGA-LIHC cohorts were previously digitized and obtained for this study. For the discovery (Gao *et al*.) and validation (Yang *et al*.) cohorts, H&E slides from formalin-fixed, paraffin-embedded blocks adjacent to molecular samples were digitized using a KF-PRO-120 scanner. This study was conducted in accordance with the Declaration of Helsinki and received ethical approval from the Ethics Committee of Zhongshan Hospital, Fudan University (B2024-262). Informed consent was obtained from all patients participating in the study.

### Data Preprocessing and Feature Extraction

For all cohorts, H&E-stained WSIs underwent a standardized digital preprocessing pipeline. In cases with multiple slides per patient, the single WSI containing the largest tumour area was selected to best capture intra-tumoral heterogeneity. Tumour regions were first automatically delineated using a previously validated classification model^29^. These regions were then systematically tiled at 10× magnification, and a pre-trained pathology foundation model, UNI^21^, was employed to extract deep feature representations from each tile for subsequent discovery and analysis.

### Morphological Landscape Mapping (MLM) Framework

To systematically decode ITH, the MLM framework employed a three-stage, multi-scale clustering strategy (Fig. 1B): (1) Data Purification: A micro-tile clustering step performed critical data purification by selecting only tiles highly enriched with tumour components. This ensured that subsequent analyses focused specifically on neoplastic morphology, minimizing confounding signals from the stromal microenvironment. (2) Intra-slide Representative Sampling: To capture the full morphological diversity within each WSI while ensuring computational feasibility for cohort-level analysis, we performed intra-slide clustering to identify local subregions, guiding a representative sampling strategy. (3) Cross-Patient Phenotype Discovery: These representative tiles were then clustered at the cohort level using a novel similarity metric based on Optimal Transport (OT), which robustly identified 16 recurrent morphological phenotypes. Finally, an OT-Prototype Network (OT-ProtoNet) classifier was developed to generalize this morphological lexicon, enabling the mapping and quantification of these 16 phenotypes across all 1,448 WSIs.

### LCM In Situ Proteomic Analysis

To define the *in situ* molecular programs, we performed map-guided LCM coupled with deep spatial proteomics in Gao *et al*. cohort (Fig. 1C). DIA-NN was used to process diaPASEF data in library-free mode, and missing values were imputed using the KNN algorithm. To identify the molecular signature of each morphological phenotypes, we performed paired differential expression analysis using the R package limma. Each phenotypic region was compared against a patient-specific internal control composed of the pooled “other” regions from the same specimen. Critically, we employed the duplicateCorrelation function to account for the inherent correlation between multiple samples from the same patient, appropriately modelling our experimental design. This analysis generated a complete, ranked list of proteins for each phenotype, which served as the primary input for Gene Set Enrichment Analysis (GSEA) against the MSigDB Hallmark gene set.

### Statistical Analysis

To establish the biological and clinical relevance of the discovered morphological phenotypes, we conducted a multi-faceted analysis. Using the deeply annotated Gao *et al*. cohort, we assessed the associations between morphological phenotype abundance (as a continuous variable) and established clinicopathological classifications (e.g., WHO subtypes^16^, tumour differentiation grade, vascular invasion), as well as clinical features (e.g., AFP levels), primarily using the Wilcoxon test. We also performed correlation analyses to compare morphological abundance with paired bulk proteomic and transcriptomic data.

The prognostic significance of individual morphological phenotypes (based on their abundance) and the MSEI was evaluated using Cox proportional hazards models and Kaplan-Meier analysis (Fig. 1D). The statistical robustness of prognostic findings in the prognostic discovery cohort (Shi *et al*., n=981) was assessed. Optimal cutoffs for survival stratification were determined in Shi *et al*. cohort and subsequently applied to the validation cohorts (Gao *et al*. and TCGA-LIHC).

To validate the biological and prognostic relevance of the key biological pathways identified from the LCM-GSEA analysis, we employed a validation pipeline using the bulk proteomic cohorts (Gao *et al*. and Yang *et al*.). All statistical analyses were performed using R (v4.2.1). A P-value < 0.05 was considered statistically significant unless otherwise specified.

## Results

### MLM Reveals 16 Recurrent Morphological Phenotypes in HCC

The MLM framework employed a hierarchical strategy to systematically decode HCC heterogeneity (Fig. 1B). First, to ensure biological specificity, a micro-tile classifier effectively distilled tumour-enriched regions from the surrounding microenvironment (balanced accuracy: 0.927; 95% CI: 0.924-0.930), minimizing confounding stromal tissues. Subsequently, intra-slide partitioning enabled a representative sampling strategy that reduced data volume by >96% while preserving rare, less abundant phenotypes (Fig. 2A-B). This process confirmed that the pathology foundation model yields stable representations across both cellular and tissue scales.

**Figure 2.**
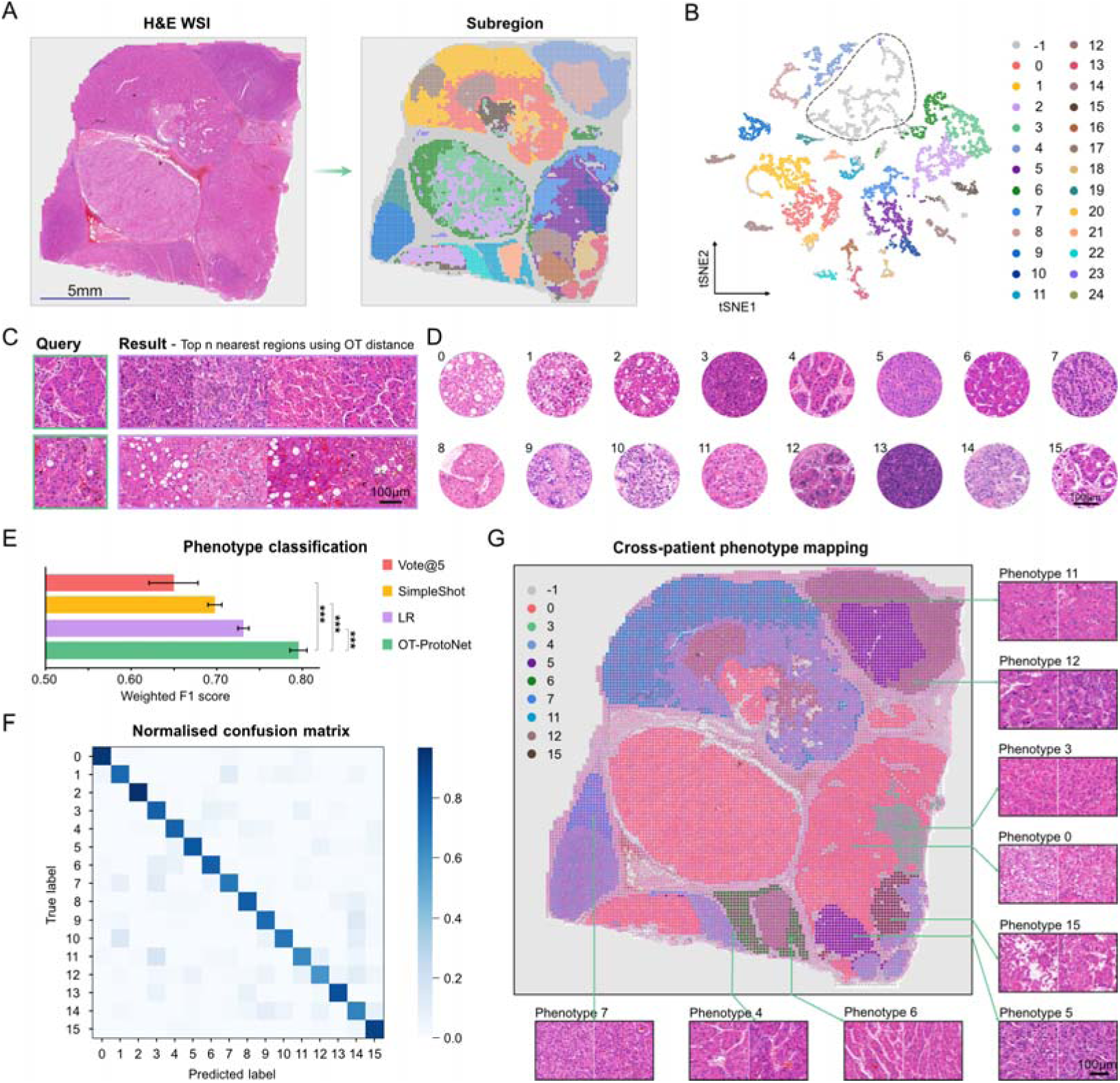
Morphological Landscape Mapping (MLM) framework for annotation-free phenotype discovery and classification in HCC. (A) Intra-slide mapping of clustering categories on a representative WSI. (B) t-SNE embedding of class-token features for all tiles in (A); each point represents a tile and is coloured by its intra-slide clustering category, whereas grey points (labelled -1 and outlined by the dashed contour) correspond to tiles with low tumour components (TC) that were not assigned to any intra-slide cluster. (C) Region-level retrieval using optimal-transport (OT) distance, ranking regions most similar to the query across different WSIs. (D) Representative morphological phenotypes identified from cross-patient clustering. (E) Performance comparison of methods for classifying 16 cross-patient phenotypes using weighted F1 score: Voting Method (Vote@5), SimpleShot, Logistic Regression (LR), and OT-ProtoNet. (F) Confusion matrix for OT-ProtoNet classification; diagonal values shaded in blue. (G) Spatial mapping of global phenotypes within each WSI using the OT-ProtoNet classifier, with representative tiles shown for each phenotype. Error bars in (E) denote 95% CIs from 25 cross-validation; P values (***, P < 0.001) calculated by paired t-test.

To identify population-level archetypes, we utilized an OT-based similarity metric. This metric significantly outperformed standard methods (e.g., Euclidean distance) in discriminating histological subtypes (Majority Vote Accuracy [MVAcc]@5: 0.954; 95% CI: 0.931-0.972; P < 0.05) by comprehensively capturing fine-grained, cell-level feature distributions, thereby preserving the local cellular diversity often lost with global aggregation methods. Qualitative validation via region retrieval confirmed the metric’s efficacy, where top-ranked matches for query regions consistently exhibited morphologically analogous patterns across diverse patients (Fig. 2C). Using this metric for cross-patient clustering, we distilled the diverse tumour landscape into 16 recurrent morphological phenotypes (Fig. 2D).

To contextualize these computationally derived patterns within diagnostic pathology, two attending pathologists provided morphological summary of the identified phenotypes. The phenotypes were characterized as follows: steatosis changes (phenotype 0); varying degrees of clear cell change (phenotypes 1 and 7); pseudoglandular architecture (phenotype 2); cytoplasm-rich cells in thin trabecular arrangements with well-differentiated features (phenotype 3) or moderate atypia (phenotype 11); normal liver-like plate architecture with single-cell-thick trabeculae (phenotype 6); pseudovascular structures (phenotype 4); distinct intercellular spaces (phenotype 5); thick trabecular arrangements (phenotype 8); abundant intratumoural stroma (phenotype 9); atypical clear cell changes with nuclear atypia (phenotype 10); nuclear pleomorphism (phenotype 12); compact cellular arrangement (phenotype 13); nuclear atypia with an increased nuclear-to-cytoplasmic ratio (phenotype 14); and loss of cell adhesion (phenotype 15). Quantitative analysis, performed by re-purposing the micro-tile classifier to quantify the pixel-wise tumour proportion within each tile, confirmed the neoplastic purity of these phenotypes, with 15 of 16 containing minimal non-neoplastic components (on average <5% stromal and/or immune-associated area).

To apply this morphological lexicon at scale, we trained an OT-Prototype Network classifier on an expanded dataset that combined expert-curated tiles from the discovery (Gao *et al*.) cohort with additional pathologist-calibrated tiles from Shi *et al*. and TCGA-LIHC cohorts. The classifier achieved high performance on the test set according to our primary metric (weighted F1 score: 0.795; 95% CI: 0.785-0.806; P < 0.001 vs. benchmark methods; Fig. 2E-F), enabling generation of comprehensive HCC morphological maps for all 1,448 cases (Fig. 2G). These maps revealed marked inter-patient heterogeneity: individual tumours harboured 1-11 distinct phenotypes with abundances ranging from 0.86% to 100%; notably, over 70.4% of patients presented with a multi-phenotypic composition. Analysis of phenotype co-occurrence further identified significant negative correlations between several phenotypes—for example, phenotype 3 was largely mutually exclusive with phenotype 14—suggesting that they represent distinct evolutionary trajectories.

### Morphological Phenotypes Associate with Clinicopathological Features and Patient Outcomes

Having defined 16 recurrent phenotypes, we assessed their prognostic significance in Shi *et al*. cohort (n = 981). Univariate analyses revealed that 14 phenotypes were significantly associated with overall survival (OS) (P < 0.05). Specifically, seven phenotypes (0, 1, 2, 3, 6, 7, 11) were indicative of favourable prognosis, whereas the other seven (4, 5, 8, 9, 10, 14, 15) were linked to poor outcomes (Fig. 3A). Notably, Phenotypes 3, 14, and 15 emerged as the most robust prognostic indicators, achieving consistent statistical significance across all three independent cohorts. For the remaining phenotypes, the direction of risk association remained consistent across cohorts, although statistical significance varied, likely due to differences in cohort size and baseline characteristics.

**Figure 3.**
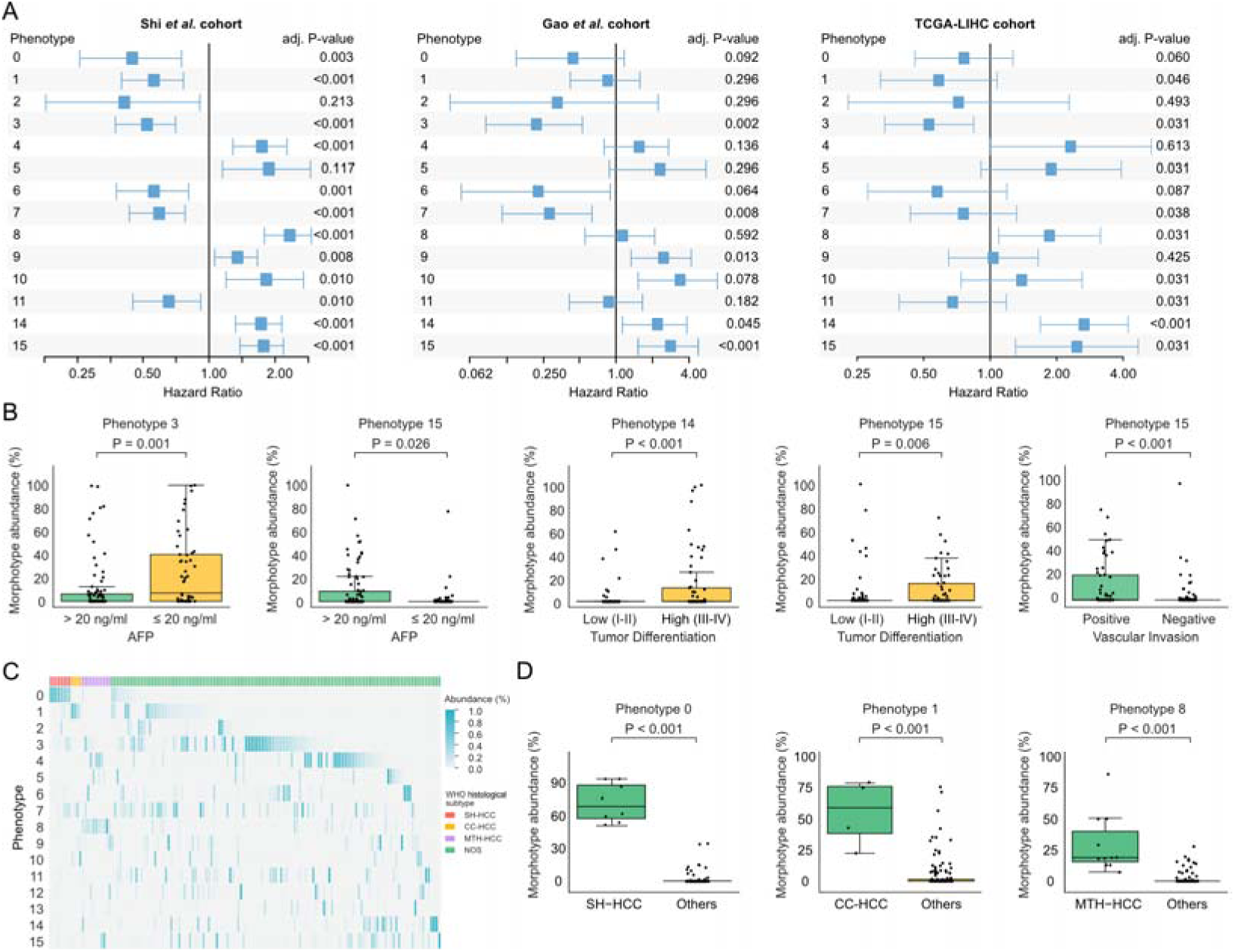
Prognostic impact and clinicopathological associations of morphological phenotypes. (A) Prognostic value of morphological phenotypes across three HCC cohorts. Forest plots showing the HR for overall survival associated with the abundance of each phenotype in Shi *et al*. cohort (n=981), Gao *et al*. cohort (n=146), and TCGA-LIHC cohort (n=274). Blue squares denote HR estimates from univariate Cox proportional-hazards models; horizontal whiskers represent 95% confidence intervals. The vertical line marks HR = 1 (no effect). Two-sided Wald P values from the Cox models were adjusted for multiple testing using the Benjamini-Hochberg false discovery rate (FDR) procedure. (B) Association with clinical features. Box plots and scatter plots illustrating the relationship between phenotype abundance and key clinical parameters in Gao *et al*. cohort. (C) Phenotype composition across WHO histological subtypes. Heatmap showing phenotype abundance (%) of the 16 MLM-defined phenotypes across individual tumours in the Gao *et al*. cohort, grouped by WHO histological subtype. Rows represent phenotypes and columns represent individual tumours; colour intensity indicates phenotype abundance (%). (D) Subtype-associated phenotypes. Box plots comparing the abundance of representative phenotypes enriched in specific WHO subtypes (Phenotype 0 for SH-HCC, Phenotype 1 for clear cell HCC, and Phenotype 8 for MTM-HCC) versus all remaining cases (“Others”). P values in (B) and (D) were calculated using two-sided Wilcoxon tests. SH-HCC, Steatohepatitic; CC-HCC, Clear Cell; MTM-HCC, Macrotrabecular-Massive; NOS, not otherwise specified.

To understand the clinicopathological basis for these survival trends, we assessed associations between phenotypes and established clinical features using the deeply annotated Gao *et al*. cohort (Fig. 3B). We observed a clear correspondence between morphological aggression and clinical progression. Specifically, high-risk Phenotypes 14 and 15 were significantly enriched in poorly differentiated tumours (Edmondson-Steiner grade III-IV). Phenotype 15 exhibited a broadly aggressive profile, showing strong correlations with both vascular invasion and elevated serum AFP levels (> 20 ng/ml). Conversely, the favourable Phenotype 3 was significantly enriched in patients with low serum AFP levels (≤ 20 ng/ml) (all P < 0.05).

Expanding this validation to histological classification, we observed striking concordance between computational phenotypes and WHO subtypes (Fig. 3C-D). Specifically, Phenotype 8 was significantly enriched in Macrotrabecular-Massive HCC, while Phenotypes 0 and 1 emerged as the predominant constituents of Steatohepatitic and Clear Cell subtypes, respectively. Crucially, this analysis decoded the latent heterogeneity of the NOS category—which constituted 84.25% of cases in Gao *et al*. cohort. In contrast to the structurally uniform specific subtypes, NOS tumours manifested as complex phenotypic mixtures, frequently composed of multiple distinct phenotypes with varying abundances.

### Morphological phenotypes bridge routine histology with proteomic subtypes

We next investigated the molecular underpinnings of these morphological patterns. Using bulk multi-omics data from Gao *et al*. cohort, we identified extensive associations between phenotype abundance and molecular profiles (Fig. 4A). In total, 7,795 proteins and 7,986 transcripts were significantly correlated with specific phenotypes (P < 0.05). Comparisons of correlation strengths revealed that 9 of the 16 phenotypes exhibited significantly stronger associations with the proteome than the transcriptome, while another 5 showed no significant difference. This dominance suggests that morphological changes more directly reflect the functional proteome than the transcriptome.

**Figure 4.**
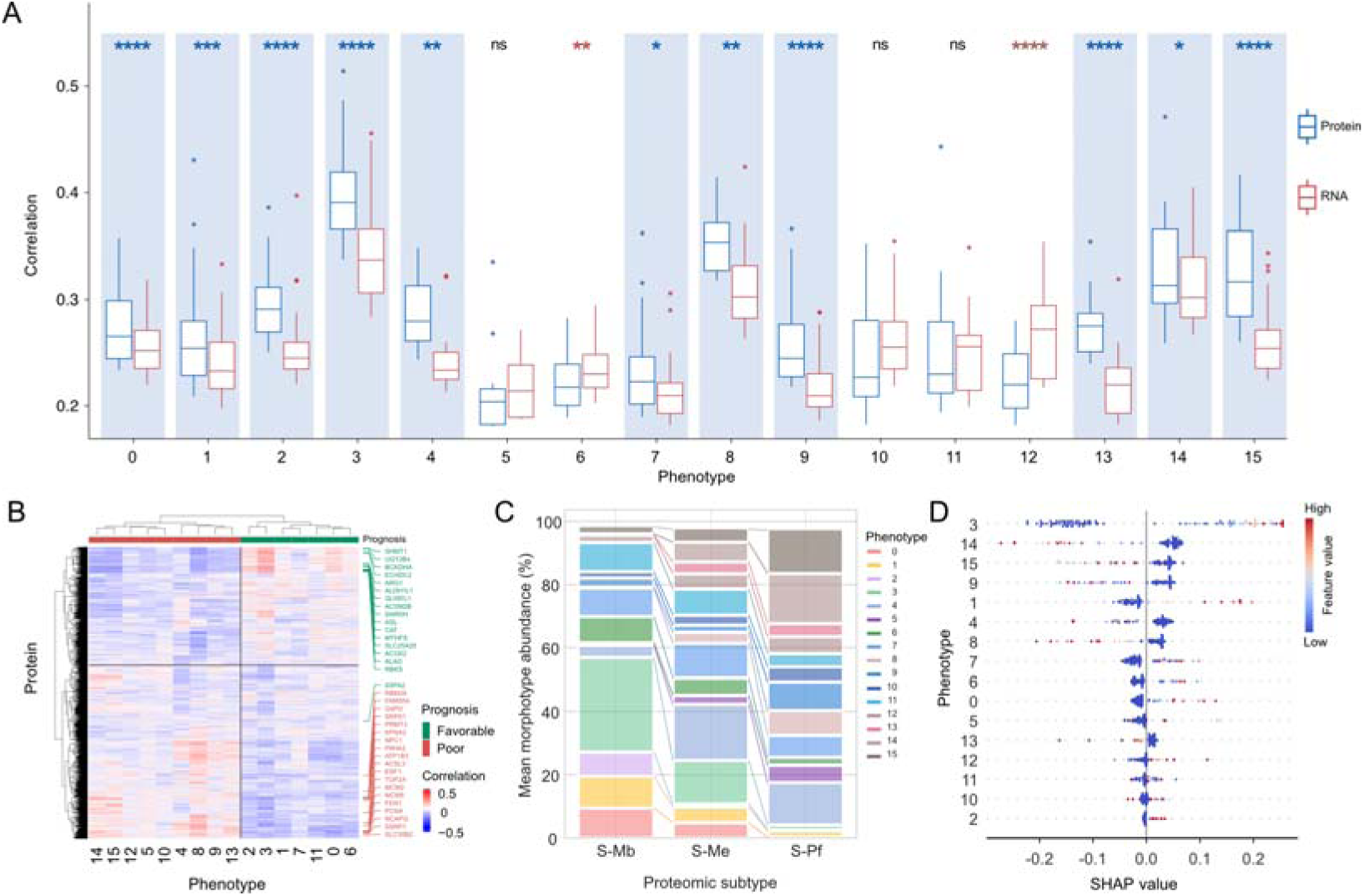
Bulk proteomic and transcriptomic associations of morphological phenotypes in Gao *et al*. cohort. (A) Boxplots of Pearson correlation coefficients between the abundance of each of the 16 morphological phenotypes and their top 200 most significantly correlated, shared molecules from bulk tumour proteomics (blue) and bulk transcriptomics (red). Blue shading marks phenotypes whose proteomic correlations are significantly stronger than their transcriptomic correlations. Statistical significance was assessed using a two-sided Wilcoxon test (* P<0.05, ** P<0.01, **** P<0.0001; ns, not significant). (B) Heatmap of Pearson correlations between phenotypes and significantly associated proteins from bulk tumour proteomes (P<0.05, coefficient of variation >20%, n=7,166). Columns and rows are ordered by hierarchical clustering. Right annotations highlight previously reported prognostic proteins (green, favourable prognosis; red, poor prognosis). (C) Stacked bar charts showing the mean phenotype composition across the three established bulk proteomic subtypes (S-Mb, metabolic; S-Me, microenvironment; S-Pf, proliferative). (D) SHAP (SHapley Additive exPlanations) summary plot for the classifier distinguishing S-Pf from S-Mb tumours based on phenotype abundances. Only samples from these two bulk proteomic subtypes were included. Each point represents a sample-phenotype pair; SHAP values on the x-axis indicate the contribution of phenotype abundance to the predicted class, with colours denoting normalized phenotype abundance (red, higher; blue, lower). Phenotypes are ordered by overall importance (top to bottom).

Hierarchical clustering of the highly variable proteins across Gao *et al*. cohort (coefficient of variation >20%, n = 7,166) revealed two major protein clusters that aligned with prognostic risk (Fig. 4B). Remarkably, our findings showed high concordance with the molecular signatures reported by Gao et al. Specifically, 16 of the 20 tumour-specific proteins associated with favourable OS (hazard ratio [HR] < 1)^32^ were enriched in the low-risk phenotype group (Phenotypes 0, 1, 2, 3, 6, 7, 11), whereas 19 of the 20 high-risk proteins were enriched in the poor-prognosis group (Phenotypes 4, 5, 8, 9, 10, 12, 13, 14, 15). These molecular associations serve to reinforce the prognostic trends independently observed in *Shi et al.* cohort. (Fig. 3A).

The strong coupling between morphology and molecular state prompted us to assess whether phenotype abundance could predict established HCC proteogenomic subtypes^32^. Phenotypes showed robust correlations with specific subtypes: Phenotype 3 predominated in the metabolic (S-Mb) subtype, whereas Phenotypes 14 and 15 were most abundant in the proliferative (S-Pf) subtype (Fig. 4C). Leveraging these morphological features, a random forest classifier distinguished S-Mb from S-Pf tumours with high accuracy (AUC = 0.910; 95% CI: 0.804-0.990). SHAP-based interpretability analysis identified Phenotypes 3, 14, and 15 as the most influential features (Fig. 4D). Expanding the model to include the microenvironment-driven S-Me subtype yielded more modest performance (macro-AUC: 0.7359; 95% CI: 0.7325-0.7393). This is likely attributable to the fact that the S-Me subtype is defined primarily by stromal rather than neoplastic features. Nevertheless, these results indicate that MLM-derived phenotypes can serve as effective, low-cost surrogates for tumour molecular subtyping.

### Deep Proteomics Define Morpho-Molecular Identities of Favourable and Aggressive Phenotypes

Given the finding that morphology is tightly coupled with the proteome, we performed phenotype-map-guided LCM to characterize the molecular context of individual phenotypes directly *in situ*. We micro-dissected 96 regions of interest (ROIs) from WSIs of the Gao *et al.* cohort and analysed them by mass spectrometry, yielding a deep dataset of 8,160 unique proteins. Principal component analysis (PCA) (Fig. 5A), paired differential expression, and functional enrichment analysis collectively confirmed that these distinct visual phenotypes are underpinned by specific, biologically coherent proteomic programs. Notably, the phenotypes that most consistently stratified survival and dominated morphology-based proteomic subtyping (Phenotypes 3, 14, and 15; Fig. 3A, 4D) also exhibited the most distinct and biologically coherent pathway-level programs *in situ*, providing a mechanistic rationale for their opposing clinical associations. Phenotype 3, visually characterized by well-differentiated trabeculae, was functionally defined by significant enrichment of the Xenobiotic Metabolism pathway (P = 0.002, Fig. 5B-C). This signature was driven by the high expression of key metabolic enzymes such as CES2, GSTO1, and PARK7, functionally recapitulating a “hepatocyte-like” state with preserved metabolic machinery—a hallmark of favourable prognosis in HCC^32^.

**Figure 5.**
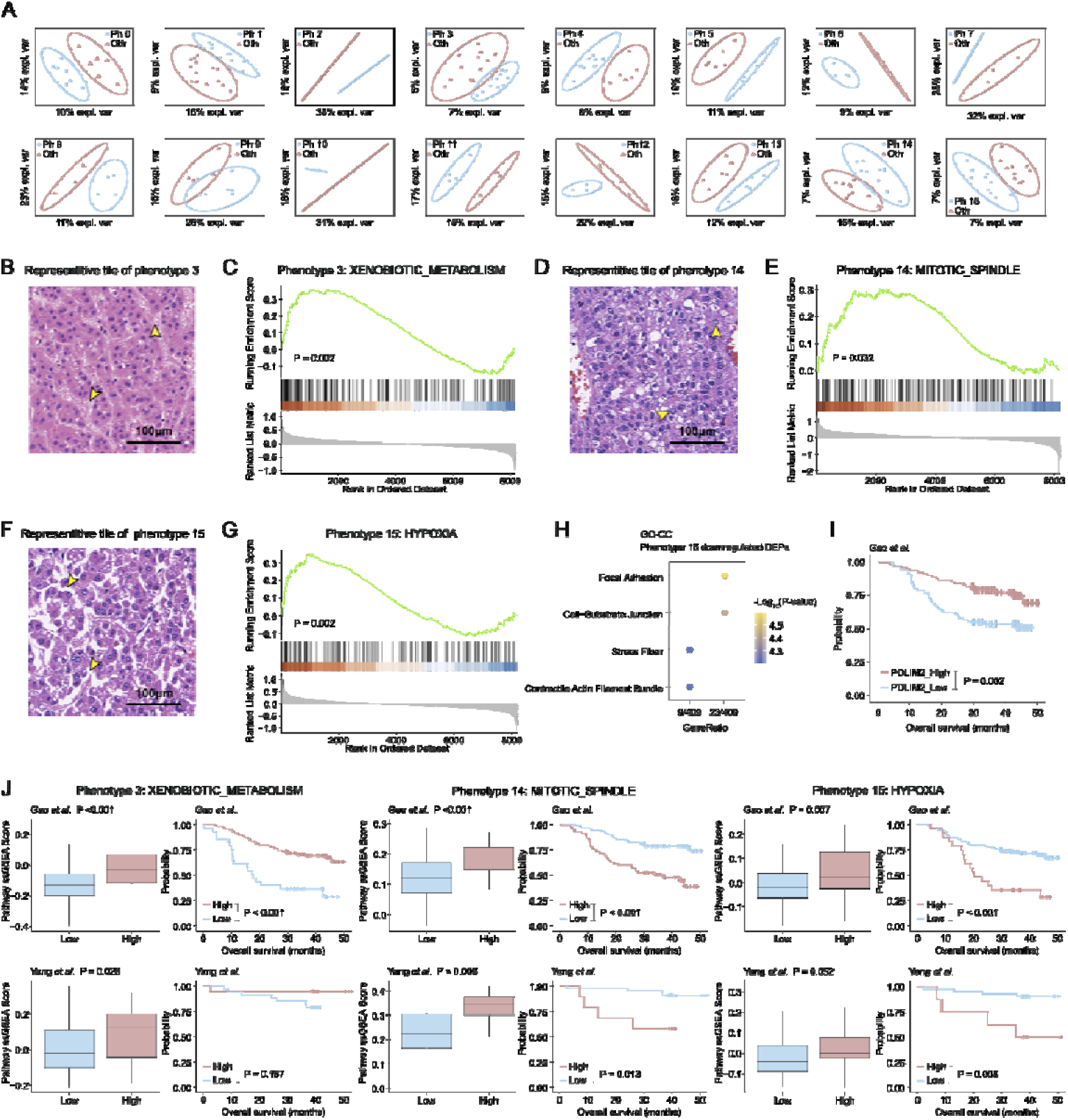
Deep spatial proteomics characterization of morphological phenotypes. (A) Principal component analysis (PCA) plot showing the distribution of proteomic profiles from microdissected phenotypic regions (Ph) and patient-matched internal controls (Oth). (B-H) Deep *in situ* proteomic analysis. (B-C) Phenotype 3: (B) Representative H&E tile. (C) Gene Set Enrichment Analysis (GSEA) plot showing enrichment of the Xenobiotic Metabolism pathway using the MSigDB Hallmark gene set. (D-E) Phenotype 14: (D) Representative H&E tile. (E) GSEA plot showing enrichment of the Mitotic Spindle pathway (Hallmark gene set). (F-H) Phenotype 15: (F) Representative H&E tile. (G) GSEA plot showing enrichment of the Hypoxia pathway (Hallmark gene set). (H) Dot plot of Gene Ontology (GO) Cellular Component enrichment analysis for significantly downregulated proteins, highlighting terms related to focal adhesion and cell-substrate junctions. (I) Prognostic analysis of PDLIM2. Kaplan-Meier curve for overall survival stratified by PDLIM2 protein expression levels in Gao *et al*. cohort. (J) Multi-cohort validation. Evaluation of the identified molecular signatures in bulk tissues from the Gao *et al*. (discovery) and Yang *et al*. (validation) cohorts. Left panels: Boxplots displaying GSVA scores for the indicated Hallmark pathways across samples with low vs. high abundance of the corresponding phenotypes. Right panels: Kaplan-Meier survival curves stratified by the GSVA scores of the respective pathways (High vs. Low). P values were calculated using Wilcoxon rank-sum test for boxplots and Log-rank test for survival analysis.

In stark contrast, the aggressive phenotypes displayed distinct mechanisms of malignancy. Phenotype 14, marked by high cellular density, showed robust activation of the Mitotic Spindle pathway (P = 0.032, Fig. 5D-E), evidenced by the upregulation of proliferation regulators including TOP2A and CDC27. More strikingly, Phenotype 15—visually defined by a “loss of cell adhesion” architecture—demonstrated remarkable concordance between form and function. Its proteomic profile was characterized by strong induction of Hypoxia signalling (P = 0.002, Fig. 5F-G) coupled with significant downregulation of Focal Adhesion components (Fig. 5H). The depletion of structural proteins such as PDLIM2 provides a mechanistic explanation for the phenotype’s fragmented morphology and its association with high metastatic potential (Fig. 5I).

To assess the generalizability of these spatially derived molecular signatures, we validated them at protein level using Gene Set Variation Analysis (GSVA) in independent cohorts. The results were highly consistent. The Xenobiotic Metabolism signature (Phenotype 3) was significantly correlated with phenotype abundance in both Gao *et al.* (P < 0.001) and Yang *et al.* (P = 0.028) cohorts, and higher pathway activity predicted better OS (P < 0.0001 and P = 0.187, respectively). Similarly, the Mitotic Spindle signature (Phenotype 14) correlated with abundance (Gao: P < 0.001; Yang: P = 0.006) and predicted worse survival (Gao: P < 0.001; Yang: P = 0.013). The Hypoxia signature (Phenotype 15) showed parallel trends, correlating with abundance (Gao: P = 0.007; Yang: P = 0.052) and poor prognosis (Gao: P < 0.001; Yang: P = 0.008) (Fig. 5J). These findings confirm that MLM-defined morphological phenotypes are not merely visual patterns but represent reliable spatial readouts of specific molecular programs.

### Quantifying ITH through Spatial Interactions of Morphological Phenotypes

As multiple morphological phenotypes frequently coexist within individual tumours (Fig. 2G), we hypothesized that their spatial organization contains critical prognostic information beyond their compositional abundance. Accordingly, we developed the MSEI, a metric derived from interaction graphs of phenotype distribution maps that measures the architectural disorder of the tumour ecosystem (Fig. 6A).

**Figure 6.**
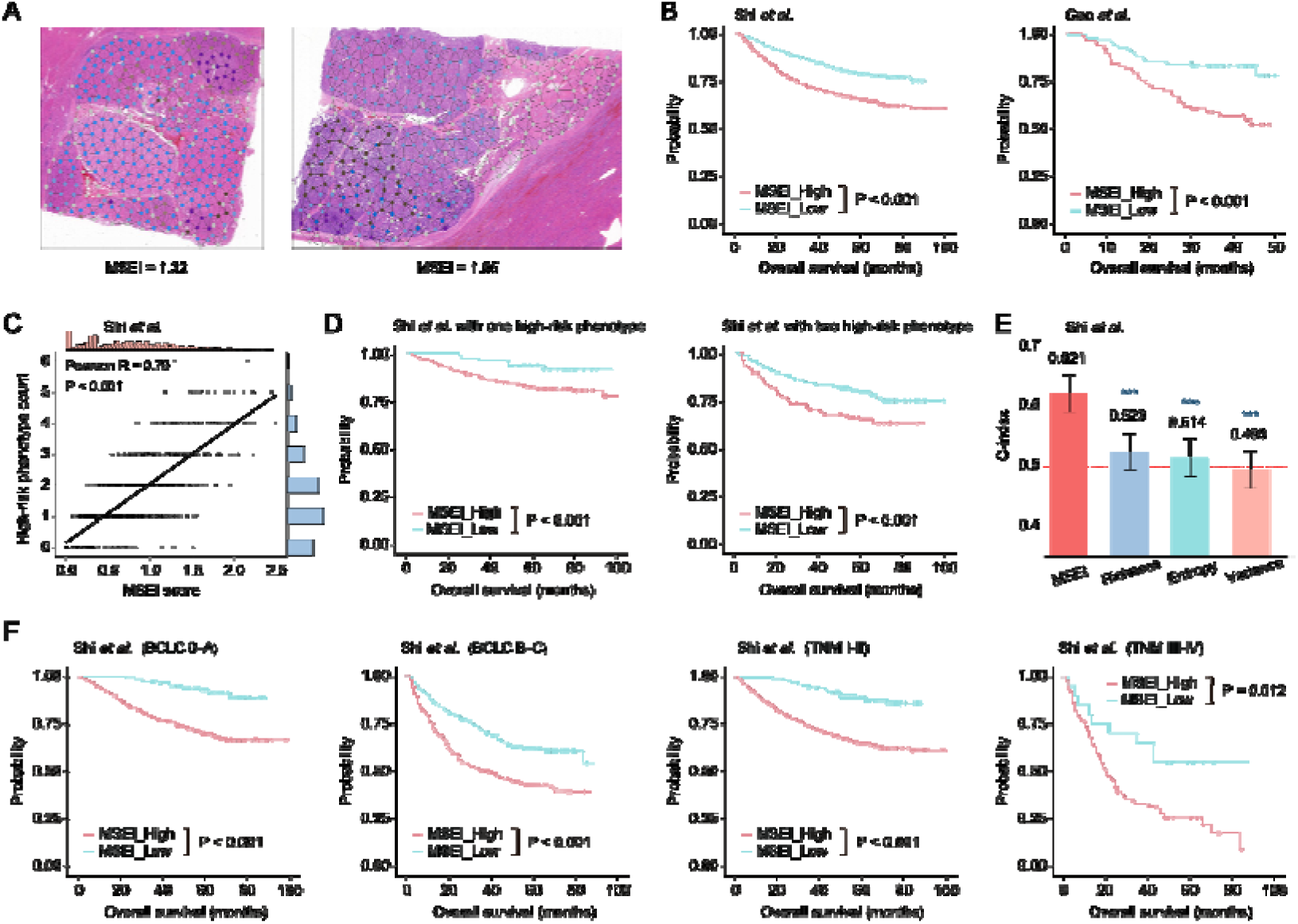
Definition of Morphotypic Spatial Entropy Index (MSEI) and its prognostic associations. (A) Construction of MSEI from whole-slide phenotype maps. Global phenotype maps are partitioned into local regions and converted into spatial interaction graphs. Nodes represent super-pixels formed by merging spatially adjacent tiles with the same phenotype. Nodes corresponding to high-risk phenotypes are colour-coded as in the global map, whereas remaining phenotypes are grouped into a single category (grey). Two representative cases with high MSEI scores are shown. (B) Kaplan-Meier curves of overall survival (OS) in Shi *et al*. and Gao *et al*. cohorts, stratified by MSEI. (C) Relationship between spatial complexity and the burden of high-risk phenotypes in Shi *et al*. cohort. Scatter plot of MSEI versus the number of unique high-risk phenotypes per tumour, with Pearson correlation coefficient and P value indicated. Marginal histograms show the distributions of MSEI values (top) and phenotype counts (right). (D) Kaplan-Meier curves of OS for patients in Shi *et al*. cohort with the same number of high-risk phenotypes, further stratified by MSEI (High vs. Low). (E) Bar plot of concordance indices (C-index) for OS prediction in Shi *et al*. cohort, comparing MSEI with compositional diversity metrics (Richness, Entropy, Variance). (F) Kaplan-Meier curves of OS in Shi *et al*. cohort showing MSEI-based stratification within clinical subgroups, including BCLC stages 0-A and B-C, and TNM stages I-II and III-IV. For all survival analyses, P values were calculated using two-sided Log-rank test. The P values (***, P < 0.001) in (E) represent the two-sided paired permutation test comparing each compositional metric against MSEI. Error bars indicate 95% CIs estimated by bootstrap resampling.

A higher MSEI indicated greater spatial complexity and robustly stratified patients into high- and low-risk groups for OS across all three cohorts (Shi *et al*. HR: 1.91, 95% CI: 1.54-2.38, P < 0.001; Gao *et al*. HR: 2.98, 95% CI: 1.70-5.22, P < 0.001; TCGA-LIHC HR: 2.41, 95% CI: 1.60-3.63, P < 0.001; Fig. 6B). Multivariate Cox proportional-hazards analysis confirmed MSEI as a strong, independent predictor of survival in Shi *et al*. cohort (HR: 2.57, 95% CI: 1.66-3.98, P < 0.001) and TCGA-LIHC cohort (HR: 2.21, 95% CI: 1.28-3.81, P = 0.005), after adjusting for key factors including BCLC/TNM stage and vascular invasion. In Gao *et al*. cohort, MSEI maintained a consistent trend toward independent prognostic value (HR: 1.79, 95% CI: 0.92-3.48, P = 0.084), though not significant due to smaller cohort size, consistent with the findings in larger cohorts.

To understand the mechanistic basis of this prognostic power, we analysed the relationship between MSEI and phenotypic composition. While MSEI scores naturally correlated with the abundance of high-risk phenotypes (Fig. 6C), this correlation does not account for its full predictive value. Crucially, among patients harbouring the exact same number of high-risk phenotypes, MSEI further stratified them into subgroups with distinct outcomes (Fig. 6D, P < 0.001). This confirms that MSEI captures architectural complexity invisible to simple counting metrics. Consequently, MSEI (C-index: 0.621, 95% CI: 0.590-0.650) demonstrated significantly superior prognostic performance compared to metrics based solely on compositional diversity (e.g., richness, entropy; all P < 0.001 vs. MSEI, Fig. 6E).

Finally, we assessed the clinical utility of MSEI in refining established staging systems. MSEI provided powerful risk sub-stratification within existing clinical stages, effectively segregating patients into high- and low-risk groups within both early (BCLC 0-A; HR: 4.34, P < 0.001) and advanced disease stages (BCLC B-C; HR: 1.85, P < 0.001), as well as across TNM stages (Fig. 6F). Furthermore, integrating MSEI with standard clinical variables (including AFP, vascular invasion, tumour size, tumour number, differentiation grade, and BCLC and TNM stages) yielded a prognostic model with a significantly higher C-index (0.741, 95% CI: 0.715-0.764) compared to these clinical variables alone (0.730, 95% CI: 0.703-0.763, P = 0.008) or the BCLC staging system (0.611, 95% CI: 0.586-0.638, P < 0.001). These data demonstrate that MSEI offers a complementary spatial dimension to current staging systems, enabling more precise risk assessment.

## Discussion

In this study, we present a comprehensive computational pathology framework that systematically decodes the morphological heterogeneity of HCC. By applying AI-driven MLM to H&E slides from 1,448 patients and integrating deep *in situ* proteomics on representative ROIs, we derived a spatially coherent phenotypic lexicon of 16 recurrent morphological phenotypes, uncovered their specific molecular underpinnings, and quantified their spatial architectural complexity. This work bridges a critical gap in precision oncology: transforming the ubiquitous, low-cost H&E slide from a qualitative diagnostic tool into a quantitative, spatially resolved, and biologically annotated map of tumour biology.

A central contribution of this work is bridging the persistent gap between visual form and molecular function. Unlike end-to-end “black box” AI models that output opaque risk scores, MLM enhances interpretability by resolving the tumour landscape into discrete phenotypes amenable to expert verification. Crucially, we confirm that these are biologically grounded morphotypes rather than arbitrary patterns. Phenotype 15 exemplifies this power: while its “loss of adhesion” features are often clinically misinterpreted as an artifact, we show it is mechanistically underpinned by downregulated cell-junction proteins (e.g., PDLIM2) and hypoxia signalling. This highlights MLM’s capacity to reveal biological insights often overlooked in routine diagnosis. The observation of analogous aggressive morphologies in other cancers by Wang *et al*.^23^ further underscores the broader significance of this phenotype in oncology. Conversely, Phenotype 3 (well-differentiated trabeculae) reflects preserved xenobiotic metabolism, suggesting that morphological patterns can serve as cost-effective surrogates for complex molecular states.

Building on this biologically validated lexicon, our framework advances diagnostic precision by shifting from dominant-pattern classification to quantitative compositional profiling. This paradigm shift effectively dismantles the ‘NOS’ wastebasket of the WHO classification, transforming a category defined by clinical ambiguity into a quantifiable spectrum of risk. By capturing sub-dominant yet prognostically significant features (e.g., Phenotype 15) that fall below traditional diagnostic thresholds, MLM recovers critical prognostic information systematically overlooked by current hierarchical classifications. This empowers pathologists to identify patients who, despite a generic diagnostic label, carry a significant burden of aggressive morphological components warranting closer clinical attention.

However, quantifying *what* constitutes the tumour provides only a compositional inventory; decoding *how* these components are spatially organized confers a higher dimension of prognostic precision. As multiple phenotypes frequently coexist, we hypothesized that their architectural configuration reflects the stability of the tumour ecosystem. Unlike existing metrics that focus primarily on compositional diversity^9,10,34–36^, the MSEI captures this architectural disorder. Its capacity to stratify risk even within early-stage disease (BCLC 0-A) suggests that high spatial complexity may serve as an independent marker of aggressive biology. By integrating this spatial dimension with compositional profiling, MLM enables pathologists to stratify risk with greater granularity, identifying high-risk patients who would otherwise be obscured by standard staging systems.

From a methodological perspective, MLM addresses the reproducibility challenge that plagues unsupervised computational pathology^30,31^. Unlike methods that focus on patient-specific variations^36^, our hierarchical strategy—combining tile-level purification with cohort-level optimal transport clustering—successfully distills population-level archetypes. This distinction is vital: clinical biomarkers must be robust across cohorts, not unique to individuals. The successful validation of our phenotypic lexicon and prognostic models across three independent cohorts (*Shi et* al., *Gao et* al., and TCGA-LIHC) underscores the generalizability of this approach.

Our study has limitations. First, the current applicability of the MLM framework is limited to HCC; extending this approach to other solid tumours will be necessary to establish its broader utility. Second, the statistical power for multivariate analysis in the multi-omics cohort was constrained by sample size, highlighting the need for larger-scale confirmatory studies. Third, as our current analysis focused on tumour cell heterogeneity, future iterations integrating the stromal and immune microenvironment would provide a more holistic view of the tumour ecosystem. Finally, coupling MLM with emerging spatial transcriptomics technologies^37,38^ represents a promising avenue to refine the molecular resolution of these morphological definitions.

In conclusion, MLM transforms routine histopathology images into quantitative, molecularly annotated maps of tumour heterogeneity. By decoding the biological and spatial logic of HCC morphology, this framework provides a scalable, cost-effective foundation for morphology-guided precision oncology, enabling more accurate prognostication and deeper biological insight from the most accessible data in cancer care.

## Data availability

The TCGA dataset used for validation is publicly available on the TCGA portal (https://portal.gdc.cancer.gov). The transcriptomic and proteomics data from Gao *et al*. and Yang *et al*. cohorts can be accessed through the National Omics Data Encyclopedia (NODE) at https://www.biosino.org/node under accession number OEP000321 and OEP002852. The processed spatial proteome data generated in this study can be accessed through the National Omics Data Encyclopedia (NODE) (https://www.biosino.org/node) with the accession number OEP00006347. Due to patient privacy and institutional review board requirements, the following data are not publicly available: the pathology images from both Gao *et al*., Shi *et al*. and Yang *et al.* cohorts, the raw spatial proteomics data from Gao *et al*. cohort, and the clinical follow-up data from Shi *et al*. cohort. Requests for access to these restricted raw data should be directed to the corresponding author.

## Acknowledgements

This work was supported by Noncommunicable Chronic Diseases-National Science and Technology Major Project (No. 2023ZD0502000); Natural Science Foundation of China (No. 82103037, 82172860, 82090053, 82130077, 82473034, 82273187 and 81961128025); Science and Technology Commission of Shanghai Municipality (No. 19XD1420700 and 23JS1410200); National Key Research and Development Program of China (2022YFC2505100); Fundamental Research Funds for the Central Universities (No. 20199215296) and High-Performance Computing Platform of Xidian University.

During the preparation of this work the authors used Google *Gemini 2.5 Pro* in order to improve readability and language. After using this service, the authors reviewed and edited the content as needed and takes full responsibility for the content of the publication.

## Author Contributions

Conceptualization: J.S., X.L., H.Z., Q.G.; Methodology: X.W., L.W., L.A., L.T., H.H.; Software: J.L., T.Z., Q.S., T.H., S.M., S.G., B.W., P.Q., Z.C., J.W., C.L.; Validation: J.H., X.Z., X.W.; Formal analysis: J.H., X.Z., X.W., H.Z., J.S., Y.C., X.P.; Investigation: J.S., Y.C., X.P., Z.Y.; Resources: J.S., Y.C., X.P., S.Z.; Data Curation: J.S., Y.C., X.P., J.H., X.Z.,S.Z.; Writing - Original Draft: X.W., J.S., Y.C., X.P.; Writing - Review & Editing: All authors; Visualization: X.W., Y.C.; Supervision: J.S., X.L., H.Z., Q.G., X.W., G.D., S.Z., L.W., L.A., L.T., H.H., J.Z., J.F.; Project administration: J.S., X.L., H.Z., Q.G.; Funding acquisition: Q.G., J.S., X.L., X.W..

